# Moderated Chain Mediation of Depression and Social Participation Between Stroke and Mortality by Socioeconomic Status

**DOI:** 10.1101/2025.05.03.25326937

**Authors:** Li Zhu, Jinhua Qian, Wenlu Shi, Zihan Geng, Siqi Yang, Tianle Wang, Lei Wang

## Abstract

**Background:** Stroke significantly increases the risk of depression and reduced social participation, particularly among middle-aged and older adults. Socioeconomic status (SES) may further shape these pathways. This study examined the mediating roles of depression and social participation, and the moderating role of SES, in the relationship between stroke and 10-year all-cause mortality in Chinese adults, while also exploring longitudinal patterns over time.

**Methods:** We analyzed data from 7,101 participants in the China Health and Retirement Longitudinal Study (CHARLS) across five waves (2011–2020). Depression was assessed via the CES-D scale, and social participation was measured based on activity frequency scores. SES was constructed from education, occupation, expenditure, and insurance. Moderated chain mediation models and generalized linear mixed models were used to evaluate pathways and temporal trends.

**Results:** Stroke was significantly associated with higher all-cause mortality. Depression partially mediated this association (β = 0.059, 95% CI [0.020, 0.023]). Although social participation alone was not a significant mediator, a sequential pathway involving depression and social participation was confirmed (β = 0.006, 95% CI [0.001, 0.012]). SES moderated both the stroke–depression and depression–social participation pathways. Notably, social participation demonstrated a protective role against post-stroke mortality when embedded within depression-related pathways—especially among low-SES individuals. Longitudinal analyses showed that the effects of stroke and depression on mortality weakened over time, while high levels of social participation consistently offered protective benefits.

**Conclusions:** Depression and social participation mediate the stroke–mortality relationship, and SES plays a critical moderating role. These findings suggest that strengthening social engagement may serve as an effective strategy to mitigate depression-related mortality among stroke survivors, particularly in low-SES populations. Tailored, SES-sensitive interventions are essential to improving survival outcomes.

## Introduction

Stroke remains the second leading cause of death and disability worldwide, marked by a high recurrence rate and a poor long-term prognosis. Beyond its immediate clinical impact, stroke imposes a substantial and lasting burden on patients, caregivers, and healthcare systems alike^1^. Among stroke survivors, long-term outcomes are often discouraging. For example, a nationwide Danish cohort study found that over 56% of stroke survivors died within 10 years of their initial stroke^2^. In China, the fact that approximately 40% of the population is now aged 45 and above underscores the urgency of this issue amid the rapid aging population^3^. This demographic shift has coincided with a marked rise in stroke incidence and mortality, particularly among middle-aged and older adults. Hence, identifying risk factors that contribute to post-stroke mortality— especially all-cause mortality—has become a critical priority for public health and clinical intervention.

A key yet under-recognized consequence of stroke is the heightened prevalence of emotional disorders, most notably depression, among survivors. Compared to general population, studies demonstrated that middle-aged and older adults are disproportionately affected with the incidence of post-stroke emotional disorders in these groups is estimated at 47.9% and 50.6%, respectively^4–6^. These findings underscore the increased susceptibility to depression in older stroke patients throughout the rehabilitation process. Moreover, longitudinal researches have shown that depressive symptoms are a significant risk factor for all-cause mortality^7^ and are associated with increased five-year mortality among elderly stroke survivors^8^. However, it is still unknown whether and to what extend post-stroke depression contributes to long-term all-cause mortality in stroke survivors. Therefore, we hypothesize that depressive symptoms mediate the relationship between stroke and all-cause mortality (**H1**).

In addition to mental health concerns, stroke-related impairments frequently results in a decline in social participation, particularly among older adults who experience physical or cognitive limitations ^9, 10^. However, accumulating evidence suggests that active social engagement serves a protective function by fostering supportive social networks and alleviating psychological distress, thereby mitigating health risks^11, 12^. Data from China Health and Retirement Longitudinal Study (CHARLS) revealed that individuals with low levels of social participation had a shorter life expectancy (12.98 years) compared to those with high levels of social participation (15.43 years)^13^. Therefore, in the present study aims to examine whether social participation may also function as a dediator between stroke and all-cause mortality (**H2**).

While depression and social participation are often treated as independent predictors of post-stroke outcomes, their interrelationship may have a synergistic effect on mortality risk. Post-stroke depression is frequently accompanied by social withdrawal, which may, in turn, intensify depressive symptoms, forming a potentially self-perpetuating cycle^14, 15^. Despite these reciprocal dynamics, few studies have explored their combined impact on mortality within an integrated framework. In this study, we therefore posit that depression and social participation operate as a sequential mediation pathway linking stroke to all-cause mortality (**H3**).

Beyond individual psychological factors, socioeconomic status (SES) may also further shape these relationships. SES encompasses key dimensions such as income, education, and occupational status, and is widely recognized as a fundamental driver of health disparities^16^. Middle-aged and older individuals with low SES face compounded vulnerabilities, limited access to healthcare, constrained social support, and elevated economic stress, factors that collectively heighten the risk of post-stroke depression, social isolation, and premature mortality^17, 18^. Prior studies have linked low SES to greater depressive symptomatology and lower levels of social participation^11, 18^. Moreover, the relationship between SES and stroke outcomes is consistently evident across both development ad developing countries^19^. However, it remains unclear whether SES not only directly predict stroke-related mortality, but also moderate the effects of depression and social participation on mortality. Accordingly, we hypothesize that SES moderates the relationship between stroke and depression (**H4**), the relationship between depression and social participation (**H5**), and the chain-mediated relationship linking stroke, depression, social participation, and all-cause mortality (**H6**).

Although the independent effects of depression, social participation, and SES on stroke outcomes are well established, their interactive mechanisms remain poorly understood. Few studies have employed an integrated longitudinal model to examine how these factors jointly influence mortality risk over time. Given the frequent co-occurrence of depression and social disengagement^20^, and the pervasive influence of socioeconomic disadvantage, this study aims to fill a critical gap by examining the chain mediation effects of depression and social participation and the moderating role of SES in the association between stroke and all-cause mortality among middle-aged and older adults.

In summary, this study proposes the following hypotheses:

**H1:** Stroke impacts all-cause mortality through the mediation of depression (*X* → *M1* → *Y*).
**H2:** Stroke impacts all-cause mortality through the mediation of social participation (*X* → *M2* → *Y*).
**H3:** Stroke impacts all-cause mortality through a chain mediation of depression and social participation (*X* → *M1* → *M2* → *Y*).
**H4:** SES moderates the relationship between stroke and depression (*M1(W)* → *M2*).
**H5:** SES moderates the relationship between depression and social participation (*M2(W)* → *M2*).
**H6:** SES moderates the chain-mediated relationship between stroke, depression, social participation, and all-cause mortality (*X(W)* → *M1* → *M2* → *Y* or *X* → *M1(W)* → *M2* → *Y*).

## Methods

### Population

This study utilized data from the China Health and Retirement Longitudinal Study (CHARLS), a nationally representative longitudinal survey designed to assess the health, economic status, social support, and psychosocial conditions of Chinese adults aged 45 years and above^21^. CHARLS was conducted by the National Institute of Development at Peking University and received ethical approval from its Institutional Review Board (IRB00001052-11015). A multistage and stratified probability sampling method was employed to ensure comprehensive coverage, encompassing 450 villages and communities across 180 counties throughout 28 provinces.

Data were collected across five waves: 2011 (baseline), 2013, 2015, 2018, and 2020. From the initial 17,708 participants, individuals under the age of 45 (n = 508) and those with incomplete baseline or follow-up data were excluded. The final analytic sample consisted of 7,101 participants. Figure 1 depicted the flowchart depicting the participant selection process.

**Figure 1.**
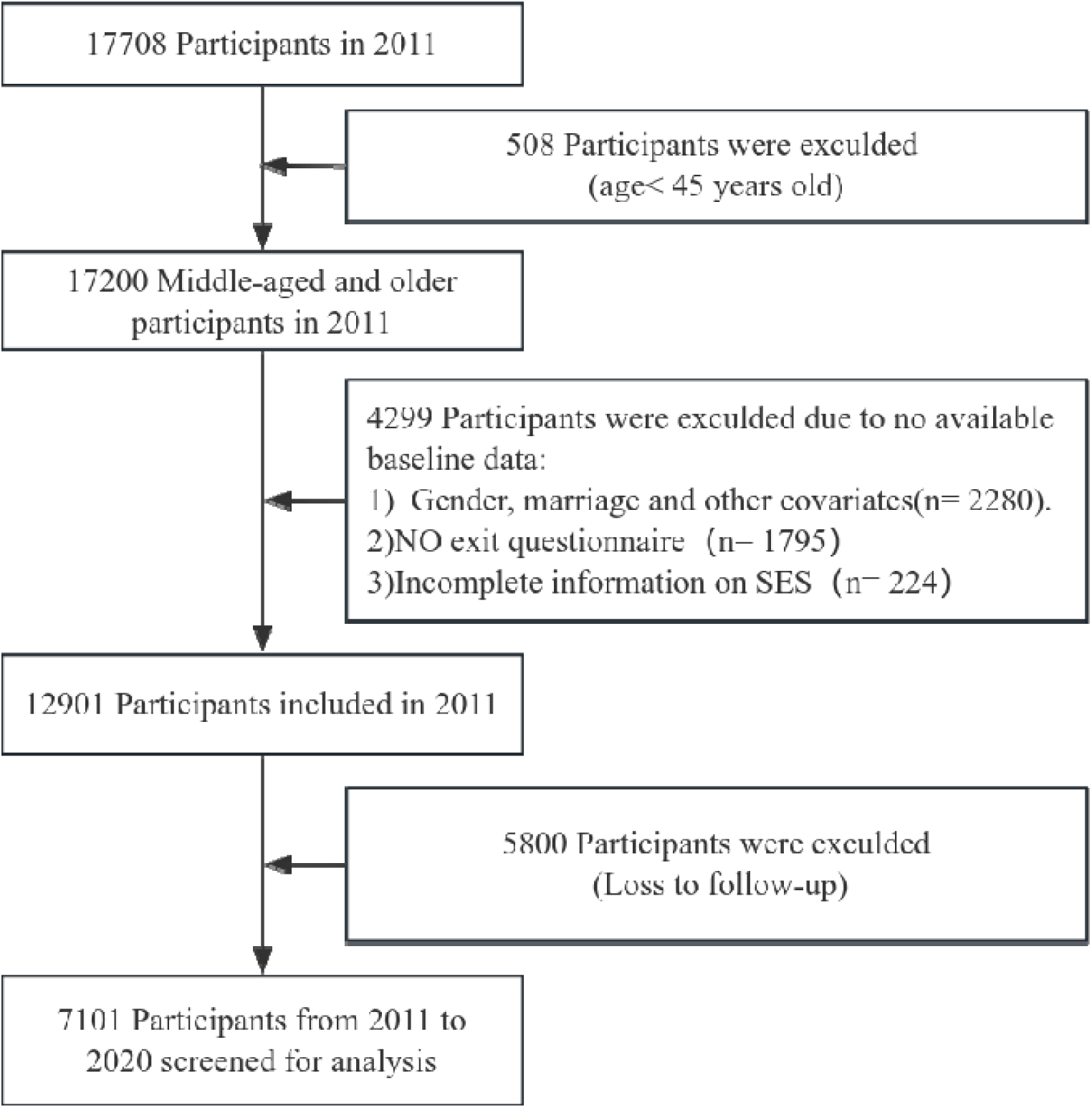
Participant selection flow chart.

### Depression

Depression was assessed using the 10-item Center for Epidemiologic Studies Depression Scale (CESD-10), a validated instrument for older Chinese adults^22^. Scores range from 0 to 30, with higher values indicating more severe depressive symptoms. The CESD-10 has demonstrated good internal reliability in previous research^23^. Item-level content is provided in Table S1.

### Social Participation

Social participation was measured by participant engagement in various activities during the past month. Frequency was self-reported on a 4-point scale including 0 (never), 1 (occasionally), 2 (almost weekly) and 3 (almost daily)^24^. Total scores ranged from 0 to 30, with higher scores referring greater social activity. Subtypes included recreational activities (items I, II, IV, V, VIII), volunteering (items III, VI, VII), and online participation (item IX), as shown in Table S2.

### Socioeconomic Status

SES was operationalized as a composite index comprising four dimensions: household expenditure per capita, education level, occupation type, and health insurance coverage. Each dimension was scored from 1 (low) to 3 (high), resulting in a total SES score ranging from 4 to 12, with higher scores indicating higher SES. Expenditure levels were categorized into tertiles according to the 33rd and 66th percentiles of the distribution:low (≤ ¥4,020), medium (¥4,020–7,541), and high (≥ ¥7,542). Education was classified into below compulsory education, high school, and university or higher^25^. Occupation was divided into unemployed, agricultural, and non-agricultural. Health insurance coverage was categorized as no insurance, public only, or both public and private coverage^26^.

### All-Cause Mortality

All-cause mortality was defined as death from any cause, recorded over the 10-year follow-up period. Mortality data were gathered through exit interviews conducted with proxies of deceased participants as part of the CHARLS tracking protocol.

### Covariates

The following variables were included as controls: gender (male or female), age group (middle-aged: 45–60; older: >60), smoking history (yes or no), marital status (married or other), and a set of chronic diseases including hypertension, diabetes, dyslipidemia, gastrointestinal disorders, chronic lung disease, cardiovascular disease, kidney disease, and psychiatric conditions.

### Statistical Analysis

All analyses were performed using IBM SPSS version 27.0. All continuous variables exhibited skewed distributions and were therefore summarized as medians and interquartile ranges (IQRs). Continuous variables were summarized as Mean ± SD, while categorical variables were described as frequencies and percentages. Between-group comparisons used independent-sample t-tests for continuous variables and chi-square or Fisher’s exact tests for categorical variables. A significance threshold of p < 0.05 was applied throughout.

Variables showing significant differences between groups were included in multivariate logistic regression models to identify predictors of all-cause mortality. Pearson correlation coefficients were computed to assess relationships between stroke, depression, social participation, SES, and mortality. Mediation and moderated mediation models were tested using the PROCESS macro for SPSS, employing bootstrapping (5,000 resamples) to estimate indirect effects with 95% confidence intervals. Generalized linear mixed models were used to explore the longitudinal effects of stroke, depression, social participation, and SES on mortality, accounting for repeated measures and within-subject variation over time.

## Result

### Descriptive Statistics and Logistic Regression

Table 1 summarize the baseline characteristics of the study sample (N = 7,101) in 2011; detailed distributions of social participation and SES subcategories are shown in Table S3. Over the 10-year follow-up period, 1,125 participants died, yielding a mortality rate of 15.84%. Significant differences were observed between the survival and mortality groups in terms of age, gender, smoking status, hypertension, diabetes, lung disease, kidney disease, heart disease, stroke history, depressive symptoms, social participation, and socioeconomic status (SES) (p < 0.05 for all).

**Table 1.**
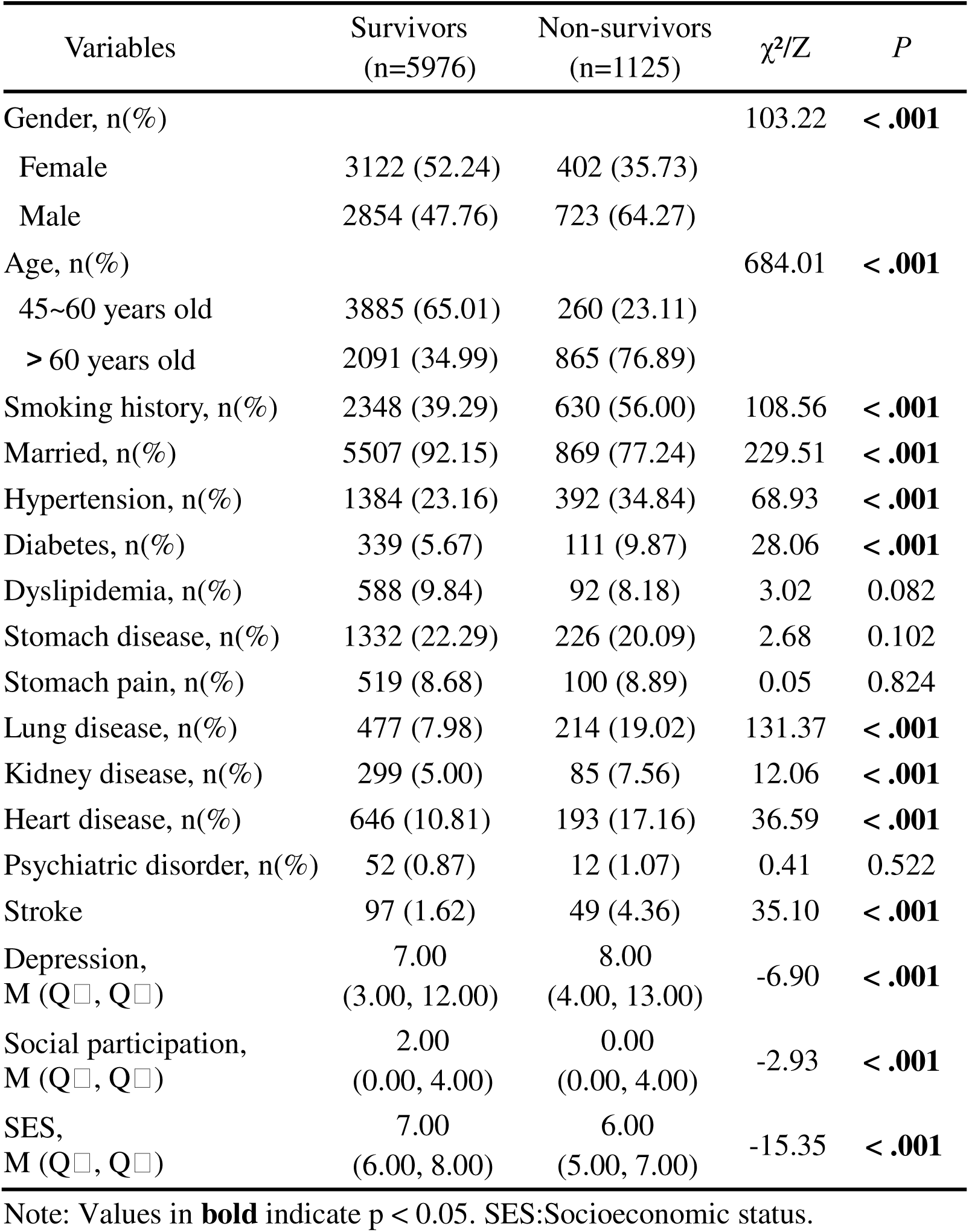
Basic characteristics and differences analysis of participants.

| Variables | Survivors<br>(n=5976) | Non-survivors<br>(n=1125) | $\chi^2/Z$ | *P* |
| --- | --- | --- | --- | --- |
| Gender, n(%) |  |  | 103.22 | < .**.001** |
| Female | 3122 (52.24) | 402 (35.73) |  |  |
| Male | 2854 (47.76) | 723 (64.27) |  |  |
| Age, n(%) |  |  | 684.01 | < .**.001** |
| 45~60 years old | 3885 (65.01) | 260 (23.11) |  |  |
| > 60 years old | 2091 (34.99) | 865 (76.89) |  |  |
| Smoking history, n(%) | 2348 (39.29) | 630 (56.00) | 108.56 | < .**.001** |
| Married, n(%) | 5507 (92.15) | 869 (77.24) | 229.51 | < .**.001** |
| Hypertension, n(%) | 1384 (23.16) | 392 (34.84) | 68.93 | < .**.001** |
| Diabetes, n(%) | 339 (5.67) | 111 (9.87) | 28.06 | < .**.001** |
| Dyslipidemia, n(%) | 588 (9.84) | 92 (8.18) | 3.02 | 0.082 |
| Stomach disease, n(%) | 1332 (22.29) | 226 (20.09) | 2.68 | 0.102 |
| Stomach pain, n(%) | 519 (8.68) | 100 (8.89) | 0.05 | 0.824 |
| Lung disease, n(%) | 477 (7.98) | 214 (19.02) | 131.37 | < .**.001** |
| Kidney disease, n(%) | 299 (5.00) | 85 (7.56) | 12.06 | < .**.001** |
| Heart disease, n(%) | 646 (10.81) | 193 (17.16) | 36.59 | < .**.001** |
| Psychiatric disorder, n(%) | 52 (0.87) | 12 (1.07) | 0.41 | 0.522 |
| Stroke | 97 (1.62) | 49 (4.36) | 35.10 | < .**.001** |
| Depression,<br>M (Q<sub>1</sub>, Q<sub>3</sub>) | 7.00<br>(3.00, 12.00) | 8.00<br>(4.00, 13.00) | -6.90 | < .**.001** |
| Social participation,<br>M (Q<sub>1</sub>, Q<sub>3</sub>) | 2.00<br>(0.00, 4.00) | 0.00<br>(0.00, 4.00) | -2.93 | < .**.001** |
| SES,<br>M (Q<sub>1</sub>, Q<sub>3</sub>) | 7.00<br>(6.00, 8.00) | 6.00<br>(5.00, 7.00) | -15.35 | < .**.001** |

Note: Values in **bold** indicate $p < 0.05$ . SES:Socioeconomic status.

Multivariate logistic regression analyses (see Table 2) revealed that stroke (p = 0.009), depression (p = 0.003), SES (p < 0.001), age (p < 0.001), gender (p < 0.001), smoking history (p = 0.009), marital status (p < 0.001), hypertension (p < 0.001), diabetes (p < 0.001), and lung disease (p < 0.001) were significantly associated with all-cause mortality. However, social participation (p = 0.223), kidney disease (p = 0.141), and heart disease (p = 0.098) were not significant predictors in the final model.

**Table 2.** Multivariate logistic regression of all-cause Mortality.

| Variables | $\beta$ | SE | Z | P | OR [95%CI] |
| --- | --- | --- | --- | --- | --- |
| Intercept | -0.17 | 0.26 | -0.66 | 0.509 | 0.84 [0.50 ~ 1.41] |
| Stroke | 0.52 | 0.20 | 2.61 | **0.009** | 1.69 [1.14 ~ 2.50] |
| Depression | 0.02 | 0.01 | 2.99 | **0.003** | 1.02 [1.01 ~ 1.03] |
| Social participation | -0.02 | 0.01 | -1.22 | 0.223 | 0.98 [0.96 ~ 1.01] |
| SES | -0.37 | 0.04 | -10.41 | < **.001** | 0.69 [0.64 ~ 0.74] |
| Man | 0.64 | 0.10 | 6.41 | < **.001** | 1.89 [1.56 ~ 2.30] |
| Age | 1.39 | 0.08 | 17.36 | < **.001** | 4.02 [3.44 ~ 4.70] |
| Smoke | 0.25 | 0.09 | 2.62 | **0.009** | 1.28 [1.06 ~ 1.54] |
| Marry | -0.83 | 0.10 | -8.68 | < **.001** | 0.43 [0.36 ~ 0.52] |
| Hypertension | 0.29 | 0.08 | 3.66 | < **.001** | 1.34 [1.15 ~ 1.57] |
| Diabetes disease | 0.56 | 0.13 | 4.28 | < **.001** | 1.75 [1.35 ~ 2.25] |
| Lung disease | 0.67 | 0.10 | 6.66 | < **.001** | 1.95 [1.60 ~ 2.37] |
| Kidney disease | 0.21 | 0.14 | 1.47 | 0.141 | 1.24 [0.93 ~ 1.64] |
| Heart disease | 0.17 | 0.10 | 1.65 | 0.098 | 1.18 [0.97 ~ 1.45] |

Note: SES: Values in **bold** indicate $p < 0.05$ . Socioeconomic status, OR: Odds Ratio, CI: Confidence Interval.

### Correlation Analysis

As illustrated in Figure 2A, stroke and depression were positively associated with all-cause mortality, while SES demonstrated a negative correlation. No significant relationship was found between stroke and social participation. These correlations provided the empirical basis for constructing a moderated chain mediation model. Figure 2B further explores the correlations between different types of social participation.

**Figure 2.**
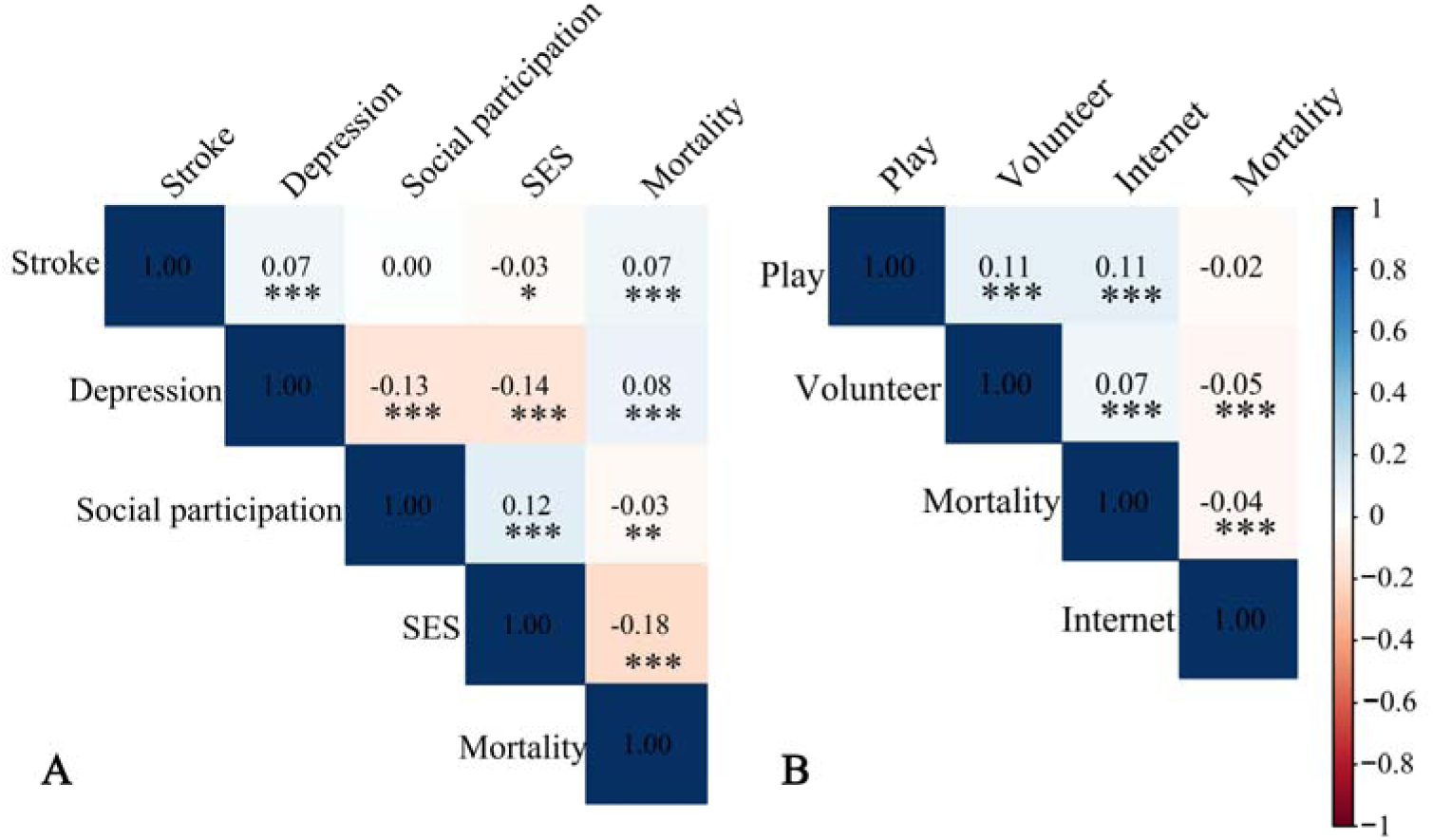
Correlation Analysis. **(A)** Pearson correlation coefficients of depression, social participation, and SES with all-cause mortality. **(B)** The correlations among different types of social participation. SES: Socioeconomic status; *p<0.05,** p < .01, ***< .001

### Chain Mediation Model

Significant predictors identified in the logistic regression were included in the mediation model to test the hypothesized indirect pathways. The path diagram is shown in Figure 3A, and detailed coefficients are reported in Table S4. Stroke was positively associated with depression (β = 0.422, p < 0.001), and depression was negatively associated with social participation (β = −0.146, p < 0.001). Depression was positively linked to all-cause mortality (β = 0.139, p < 0.001), while social participation was inversely associated with mortality (β = −0.094, p < 0.05). These findings support the presence of a chain mediation mechanism.

**Figure 3.**
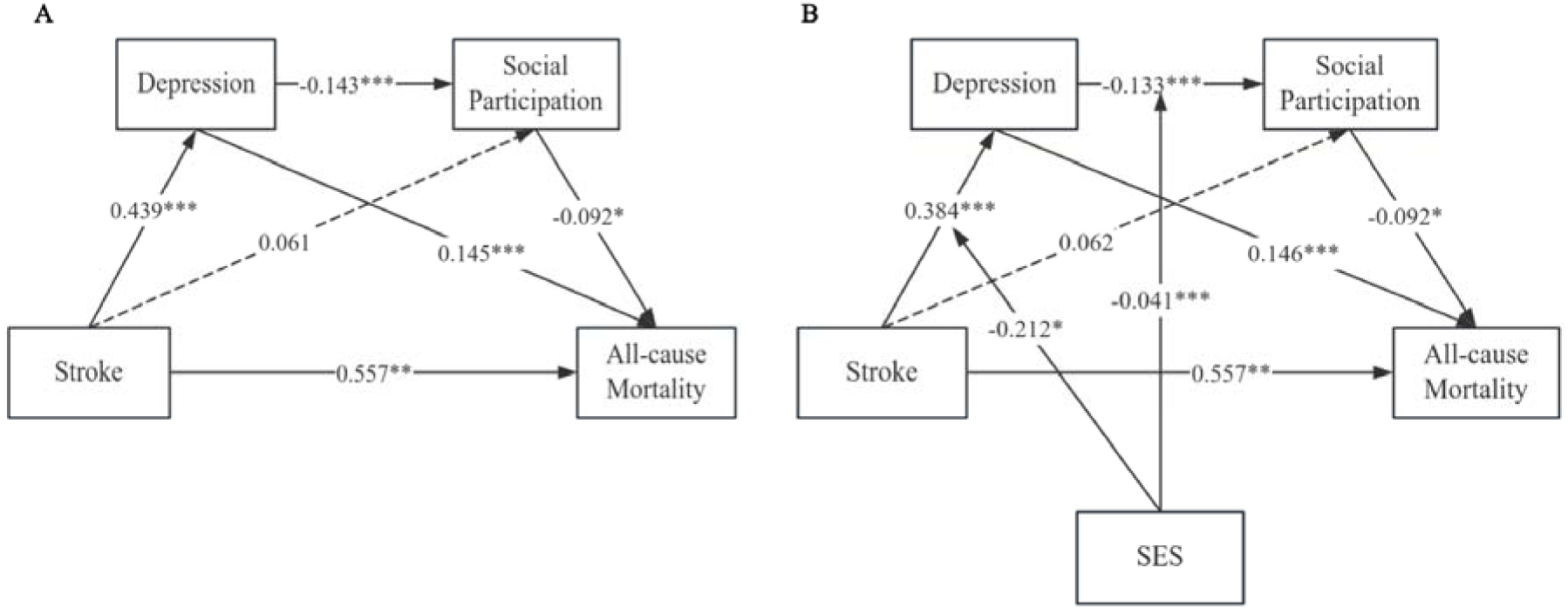
Pathways between stroke and all-cause mortality. **(A)** Depression and social participation mediated the association between stroke and all-cause mortality. **(B)** Socioeconomic status moderated the mediating effects of depression and social participation on the relationship between stroke and all-cause mortality. SES: Socioeconomic status; Solid lines: Significant; Dashed line: Not significant; *p<0.05,** p < .01, ***< .001

Bias-corrected bootstrap analyses (5,000 samples) were used to estimate indirect effects. As shown in Table S5, the indirect effect of stroke on mortality through depression (Pathway Ind1) was significant (β = 0.059, 95% CI [0.023 ∼ 0.102]), supporting Hypothesis 1 (H1). The indirect effect through social participation alone (Pathway Ind2) was not significant (β = −0.006, 95% CI [−0.027, 0.009]), as the confidence interval included zero, thereby refuting Hypothesis 2 (H2). However, the indirect effect through the sequential pathway of depression and social participation (Pathway Ind3) was significant, albeit small (β = 0.006, 95% CI [0.001, 0.012]), providing support for Hypothesis 3 (H3).

### Moderated Chain Mediation Model

To examine the moderating role of SES, PROCESS macro models 83 and 91 were applied, focusing on the chain pathway Stroke → Depression → Social Participation → Mortality. The interaction effects between SES and stroke (β = −0.243, p < 0.01), and between SES and depression (β = −0.041, p < 0.001), were both significant, supporting Hypotheses 4 and 5 (H4 and H5).

Simple slope analysis, illustrated in Figure 4, further clarifies the nature of these interactions. The moderated mediation index of SES (Table S6) on the stroke → depression → mortality pathway was −0.003 (95% CI [−0.007, −0.001]), and the index for the depression → social participation → mortality pathway was 0.002 (95% CI [0.0003, 0.004]). Since both confidence intervals excluded zero, these findings confirm a significant moderated chain mediation effect, thus supporting Hypothesis 6 (H6). The full moderated chain mediation model is presented in Figure 3B.

**Figure 4.**
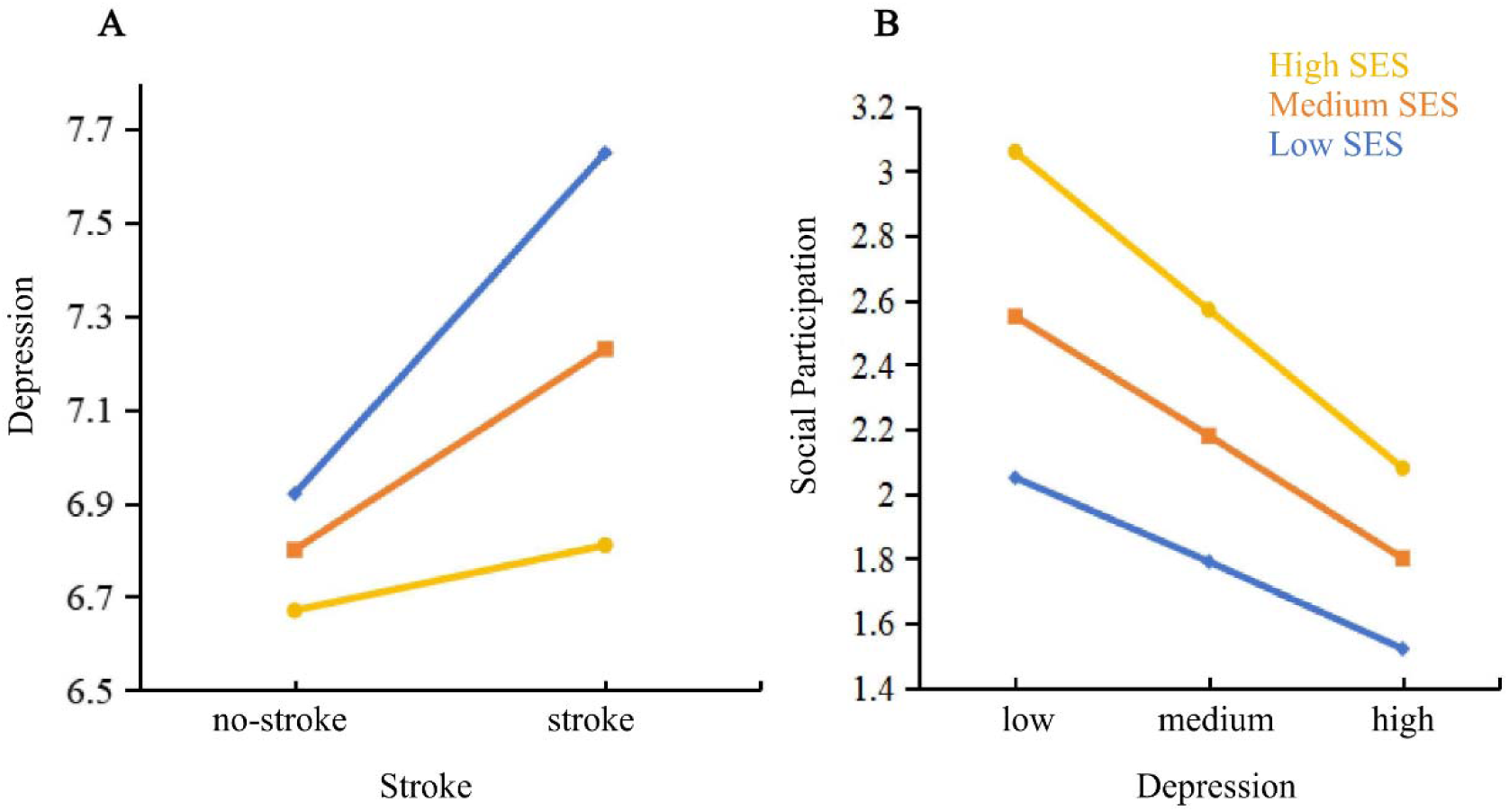
The moderating role of SES in nodes of Ind3. **(A)** The positive predictive effect of stroke on depression (standardized using Z-scores) was more pronounced among individuals with low SES. **(B)** The negative effect of depression on social participation (standardized using Z-scores) was more pronounced among individuals with high SES. SES: Socioeconomic status, Ind3: Stroke → Depression → Social Participation→ Mortality.

### Longitudinal Analysis

To assess dynamic changes in risk over time, generalized linear mixed models were used to evaluate the longitudinal relationships among stroke, depression, social participation, SES, and all-cause mortality. The results, detailed in Table S7, indicate that depression and SES remained consistent predictors of mortality across waves. Figure 5 presents subgroup analyses that demonstrate varying effects of social participation on mortality over time, accounting for temporal fluctuations and individual-level variability.

**Figure 5.**
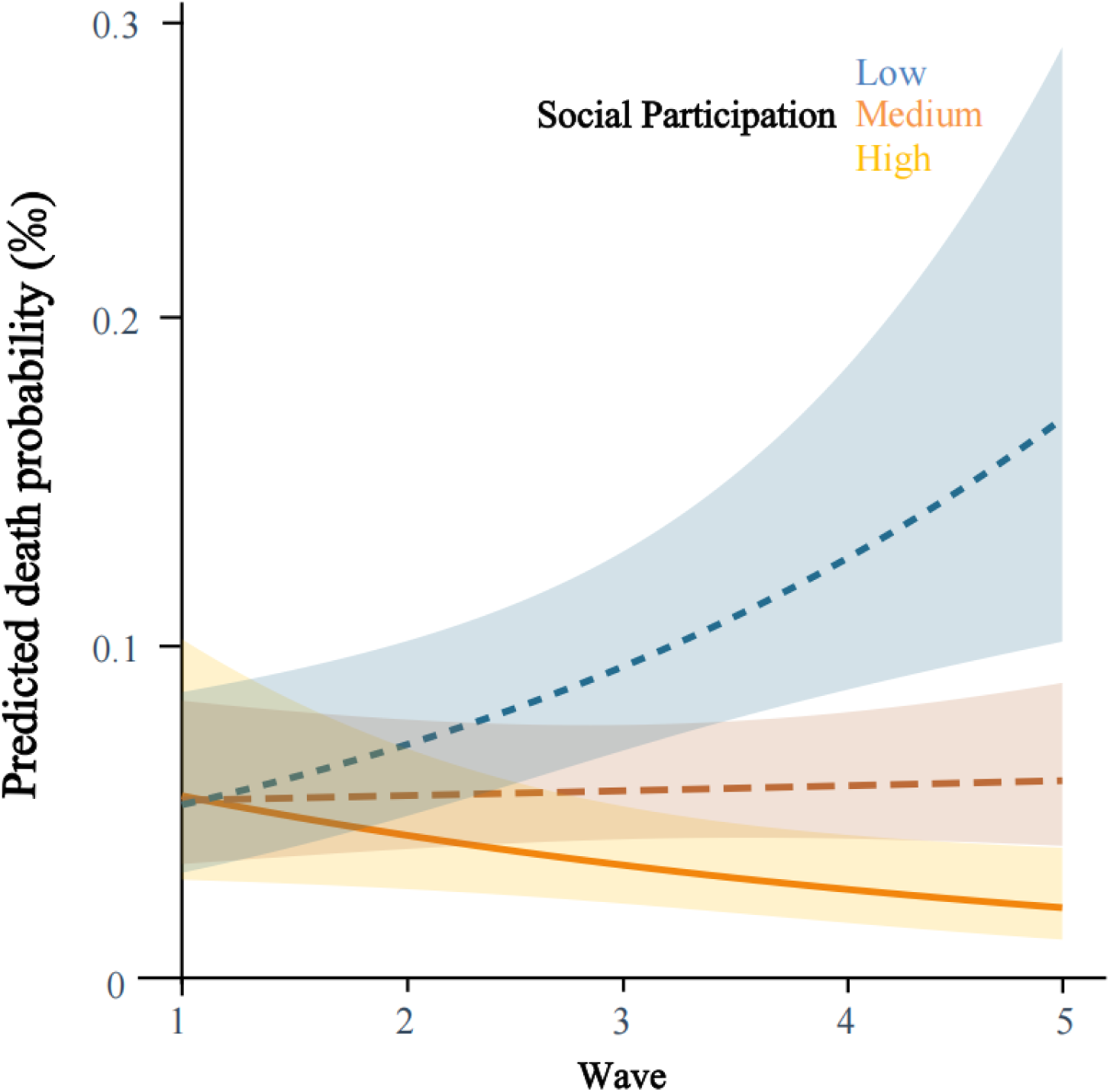
The interaction between social participation and time indicated that, over time, lower levels of social participation were associated with a progressively increasing risk of all-cause mortality, whereas higher levels of social participation were linked to a reduced risk of all-cause mortality.

## Discussion

This study investigated the complex interplay between stroke, depression, social participation, and socioeconomic status (SES) in predicting all-cause mortality among middle-aged and older adults in China. Drawing on a nationally representative longitudinal dataset and applying moderated chain mediation models, the findings reveal both direct and indirect pathways through which stroke impacts mortality risk. Notably, stroke was associated with increased all-cause mortality via partial mediation by depression (Ind1) and a sequential chain involving both depression and social participation (Ind3). Although social participation alone did not significantly mediate the stroke–mortality relationship, its protective effect was evident when coupled with depression, highlighting the importance of addressing both emotional and social well-being in post-stroke care.

The observed all-cause mortality rate of 15.84% is consistent with estimates from previous studies, such as the Chinese Longitudinal Healthy Longevity Survey^27^. Individuals in the mortality group exhibited higher depression scores and stroke prevalence, alongside lower SES and reduced social participation. Multivariate regression showed no significant direct association between social participation and mortality, suggesting that its influence may emerge through interactive or conditional pathways, especially in the presence of depression.

Approximately half of middle-aged and older stroke survivors experience depressive symptoms and social disengagement^4, 28^. The vascular depression hypothesis provides a possible explanation, attributing this vulnerability to cerebrovascular damage in brain regions responsible for emotion and cognition^6, 29^. Stroke-related neurological impairment not only leads to physical disability but also increases psychological risk— particularly depression—which, in turn, contributes to elevated mortality. Our findings align with previous research demonstrating that depression mediates cardiovascular-related mortality with similar effect sizes^30^, reinforcing that depression significantly mediates the relationship between stroke and mortality and highlighting its critical role as a clinical target in post-stroke rehabilitation^2, 31, 32^.

Interestingly, while social participation did not serve as a standalone mediator, its protective effect became salient within a chain mediation model. Stroke-induced depression leads to lower social engagement, which subsequently increases mortality risk ^28^. This finding aligns with the biopsychosocial model^33^, where emotional and behavioral states are closely interlinked. Physical limitations (e.g., aphasia, hemiplegia) can restrict social involvement, and emotional disturbances such as depression further exacerbate withdrawal^34^. Conversely, low levels of social participation can reinforce depressive symptoms, creating a feedback loop. Prior studies, including animal research, also demonstrate the exacerbating effects of social isolation on depressive symptoms^35–37^. In humans, behavioral activation therapy promoting social interaction has shown efficacy in alleviating depression, reinforcing the significance of social engagement in psychological recovery^38–40^.

Our findings also highlight the moderating role of SES in this pathway. SES negatively moderated the stroke–depression link and positively moderated the depression–social participation relationship. Individuals with lower SES were more vulnerable to depression following stroke, due to factors such as reduced access to healthcare, lower health literacy, and economic stress^4, 41, 42^. In contrast, higher SES was associated with greater resilience and better medical access^11^. However, when depression occurred among high-SES individuals, its impact on social participation was amplified. This may stem from identity concerns, as high-SES individuals often engage in more formal social activities and may experience stronger social stigma or self-imposed withdrawal when their functioning declines^43–45^.

Moreover, the chain mediation pathway itself was significantly moderated by SES. Low SES intensified the negative effects of depression and diminished social participation on mortality, whereas high SES buffered these effects^4, 19, 46^. This dual role of SES—as both a risk and protective factor—underscores the need for tailored intervention strategies that consider economic and social disparities^47^. High-SES individuals may benefit more from psychological support addressing self-identity and social reintegration, while low-SES populations may require broader access to mental health services and community-based resources^48, 49^.

Longitudinal analysis further showed that the associations among stroke, depression, social participation, and mortality are dynamic rather than static. The effects of stroke and depression on mortality declined over time, suggesting that early-stage intervention is especially critical^50, 51^. Although social participation did not display a strong fixed effect, its interaction with time revealed important variations. Individuals with higher social participation maintained lower mortality risk across time, emphasizing the cumulative benefits of social engagement^18^.

Several limitations should be acknowledged. First, inconsistencies in the measurement of social participation across survey waves—particularly in 2020—may introduce bias in cross-time comparisons, despite the applied adjustments. Second, while this study focused on all-cause mortality, it did not explore cause-specific mechanisms or account for biological pathways linking stroke, depression, and social withdrawal to mortality outcomes. Third, due to data limitations, longitudinal mediation effects were not tested, and causality cannot be definitively established from the current model structure. Finally, the findings are subject to limitations in generalizability due to the observational nature of the study and reliance on internal validation. Future research should incorporate neurobiological measures, expand to diverse populations, and apply prospective designs to validate and deepen understanding of these complex interrelationships.

In conclusion, this study demonstrates that stroke increases the risk of all-cause mortality among middle-aged and older adults, with depression and social participation acting as important mediators. SES not only influences these relationships directly but also moderates their pathways, reinforcing the importance of a multifaceted intervention approach. Social and emotional health should be considered alongside physical rehabilitation in stroke recovery programs. Efforts to promote early screening for depression and encourage meaningful social engagement can play a vital role in improving survival outcomes. Tailored interventions based on SES are crucial for ensuring equitable access to care and optimizing the long-term well-being of stroke survivors.

## Data Availability

All data produced are available online at http://charls.pku.edu.cn.

## Acknowledgments

Conceptualization, T.W., L.W.; methodology, J.Q., L.W.; investigation, J.Q.,W.S, Z.G, S,Y.; formal analysis, L.Z., J.Q.; visualization, L.Z., J.Q.; writing – original draft, L.Z., J.Q.; writing – review & editing, L.Z., J.Q., T.W., L.W.; supervision, T.W., L.W.; funding acquisition, L.Z., L.W..

## Supplemental Material

Supplemental Tables 1 – 7

## Non-standard Abbreviations and Acronyms

SES: Socioeconomic status
CHARLS: China Health and Retirement Longitudinal Survey
CI: Confidence Interval
CESD-10: 10-item Center for Epidemiologic Studies Depression Scale
IQRs: interquartile ranges
SD: Standard deviation

