## supplemental Table for "Moderated Chain Mediation of Depression and Social Participation Between Stroke and Mortality by Socioeconomic Status"

**Table S1** The 10-item Center for Epidemiological Studies Depression Scale.

| In the past week | Rarely or none  （<1 day） | Some or a little  （1-2 days ） | Occasionally or a moderate amount （3-4 days） | Most or all  （5-7 days） |
| --- | --- | --- | --- | --- |
| **(I)** I was bothered by things that don’t usually bother me | □0 | □1 | □2 | □3 |
| **(II)** I had trouble keeping my mind on what I was doing | □0 | □1 | □2 | □3 |
| **(III)** I felt depressed | □0 | □1 | □2 | □3 |
| **(IV)** I felt everything I did was an effort | □0 | □1 | □2 | □3 |
| **(V)** I felt hopeful about the future | □3 | □2 | □1 | □0 |
| **(VI)** I felt fearful | □0 | □1 | □2 | □3 |
| **(VII)** My sleep was restless | □0 | □1 | □2 | □3 |
| **(VIII)** I was happy | □3 | □2 | □1 | □0 |
| **(IX)** I felt lonely | □0 | □1 | □2 | □3 |
| **(X)** I could not get ”going” | □0 | □1 | □2 | □3 |

Note: Each item was scored based on its frequency over the past week, using a scale ranging from 0 (<1 day) to 3 ( 5–7 days). Items (V) and (VIII) were reverse-scored.

**Table S2** The social participation scale.

| In the past month | Never | Not regularly | Almost every week | Almost daily |
| --- | --- | --- | --- | --- |
| **(I)** Interacted with friends | □0 | □1 | □2 | □3 |
| **(II)** Played Ma-jong, played chess, played cards, or went to community club | □0 | □1 | □2 | □3 |
| **(III)** Provided help to family, friends, or neighbors who do not live with you | □0 | □1 | □2 | □3 |
| **(IV)** Went to a sport, social, or other kind of club | □0 | □1 | □2 | □3 |
| **(V)**Took part in a community-related organization | □0 | □1 | □2 | □3 |
| **(VI)** Done voluntary or charity work | □0 | □1 | □2 | □3 |
| **(VII)** Cared for a sick or disabled adult who does not live with you | □0 | □1 | □2 | □3 |
| **(VIII)** Attended an educational or training course | □0 | □1 | □2 | □3 |
| **(IX)** Used the Internet | □0 | □1 | □2 | □3 |
| **(X)** None of these | □0 | □1 | □2 | □3 |

Note: Each item was scored based on its frequency over the past month, using a scale ranging from 0 (<1 day) to 3 ( 5–7 days).

In the fifth wave of the survey conducted in 2020, adjustments were made due to the COVID-19 epidemic in China. Specifically:Items (III) and (VII) were merged into a new category (XI), as participants were less likely to differentiate between the two types of caregiving activities during the pandemic.For item (IX), only the occurrence of Internet use was recorded, without capturing its frequency.To ensure consistency in social participation scoring across the five survey waves, the new option (XI) in 2020 was retroactively divided into items (III) and (VII), each assigned equal values. Additionally, for Internet use in 2020, participants who reported using the Internet during the past month were assumed to engage in "almost daily" use.

**Table S3** Distribution and comparative analysis of SES and social participation subgroups in the study population.

| Variables | Survivors  (n=5976) | Non-survivors  (n=1125) | χ² | *P* |
| --- | --- | --- | --- | --- |
| Social participation |  |  |  |  |
| Paly, n(%) | 2824 (47.26) | 499 (44.36) | 3.20 | 0.074 |
| Volunteer, n(%) | 542 (9.07) | 62 (5.51) | 15.41 | **< .001** |
| Internet, n(%) | 130 (2.18) | 6 (0.53) | 13.59 | **< .001** |
| SES |  |  |  |  |
| Expenditure, n(%) |  |  | 25.12 | **< .001** |
| 1 | 2518 (42.14) | 559 (49.69) |  |  |
| 2 | 1767 (29.57) | 313 (27.82) |  |  |
| 3 | 1691 (28.30) | 253 (22.49) |  |  |
| Education level, n(%) |  |  | 45.52 | **< .001** |
| 1 | 5234 (87.58) | 1063 (94.49) |  |  |
| 2 | 655 (10.96) | 52 (4.62) |  |  |
| 3 | 87 (1.46) | 10 (0.89) |  |  |
| Health insurance, n(%) |  |  | 29.94 | **< .001** |
| 1 | 260 (4.35) | 88 (7.82) |  |  |
| 2 | 5499 (92.02) | 1013 (90.04) |  |  |
| 3 | 217 (3.63) | 24 (2.13) |  |  |
| Occupation, n(%) |  |  | 391.96 | **< .001** |
| 1 | 1296 (21.69) | 554 (49.24) |  |  |
| 2 | 3924 (65.66) | 525 (46.67) |  |  |
| 3 | 756 (12.65) | 46 (4.09) |  |  |

Note: χ²: Chi-square test, SES: Socioeconomic status.

**Table S4 Bias-corrected Bootstrap tests of direct effects between variables**

| Reult variable | Predictor variable | β | 95%CI |
| --- | --- | --- | --- |
| Depression | Stroke | 0.422 | [0.244, 0.602] |
| Social | Stroke | 0.062 | [-0.095, 0.229] |
|  | Depression | -0.146 | [-0.168, -0.124] |
| Mortality | Stroke | 0.556 | [0.162, 0.931] |
|  | Depression | 0.139 | [0.068, 0.209] |
|  | Social participation | -0.094 | [-0.170, -0.019] |

**Table S5 Bias-corrected Bootstrap tests of the chain mediation effect**

| Direct and Indirect effects | Effect | SE | 95%CI |
| --- | --- | --- | --- |
| Direct effect | 0.556 | 0.199 | [0.166, 0.945] |
| Total indirect effect | 0.059 | 0.023 | [0.016, 0.106] |
| Ind1：Stroke → Depression→ Mortality | 0.059 | 0.02 | [0.023, 0.102] |
| Ind2：Stroke → Social participation→ Mortality | -0.006 | 0.009 | [-0.027, 0.009] |
| Ind3: Stroke → Depression→ Social participation→ Mortality | 0.006 | 0.003 | [0.001, 0.012] |

**Table S6 Moderated chain mediation effects in the pathway of Ind**

| SES | Effect | SE | 95% CI |
| --- | --- | --- | --- |
| Mediating effects through Stroke and Depression | | | |
| Low | 0.008 | 0.004 | [0.002, 0.016] |
| Medium | 0.005 | 0.002 | [0.001, 0.010] |
| High | 0.002 | 0.002 | [-0.002, 0.006] |
| Total | -0.003 | 0.002 | [-0.007, -0.001] |
| Mediating effects through Depression and Social Participation | | | |
| Low | 0.004 | 0.002 | [0.001, 0.008] |
| Medium | 0.005 | 0.003 | [0.001, 0.011] |
| High | 0.007 | 0.003 | [0.002, 0.015] |
| Total | 0.002 | 0.001 | [0.0003, 0.004] |

Note: Ind3: Stroke → Depression → Social Participation → Mortality.

**Table S7 Results from generalized linear mixed models**

| Variable | estimate | SE | statistic | *P* |
| --- | --- | --- | --- | --- |
| (Intercept) | -10.204 | 0.301 | -33.876 | **< .001** |
| wave | 0.138 | 0.079 | 1.739 | 0.082 |
| Stroke | 7.032 | 0.92 | 7.642 | **< .001** |
| Depression | 1.153 | 0.18 | 6.392 | **< .001** |
| Social participation | 0.294 | 0.244 | 1.206 | 0.228 |
| SES | -0.557 | 0.227 | -2.452 | **0.014** |
| wave:Stroke | -2.491 | 0.382 | -6.517 | **< .001** |
| wave:Depression | -0.375 | 0.063 | -6.001 | **< .001** |
| wave:Social participation | -0.268 | 0.078 | -3.431 | **< .001** |
| wave:SES | 0.077 | 0.077 | 0.996 | 0.319 |

Note: Values in **bold** indicate p < 0.05. SES:Socioeconomic status
